# Moving from Total Concentrations to Measures of Harm in Grain Sold at Selected Markets of Southwest Nigeria

**DOI:** 10.1101/2022.12.18.22283634

**Authors:** Modupe Abeke Oshatunberu, Adebayo Oladimeji, Sawyerr Olawale Henry, Opasola Afolabi Olaniyan, Morufu Olalekan Raimi

## Abstract

Regardless of where you live or who you are, food safety is crucial for human health. Pesticide residues are commonly exposed to consumers in Nigeria through their food. What consequences, if any, such chemical pollutants cause to consumer health remain unclear given the presence of pesticide residues in food. To this end, the objective is to determine the concentration of the identified pesticide in grains commonly used by farmers, and which are available directly from the open markets in the Nigerian market. Pesticide residues were quantified through a multiresidue method using a varian 3800/4000 gas chromatograph mass spectrometer was used to analyze pesticide residues. The simultaneous determination of four classes of pesticides: carbamates, organochlorines, organophosphates, and pyrethroids by gas chromatography-mass spectrometry (GC/MS) method using sample preparation on QuEChERS-citrate, was developed and validated. The results frequently showed high inter- and intra-specific contamination, which makes sense given the target market and dietary diversity in the area. This study found that grains purchased from particular markets in southwest Nigeria contained numerous pesticide residues. The MRLs set by the EU or FAO/WHO or both were surpassed by 17 out of the total 27 pesticides reported in this work in at least one grain, despite the fact that there were no published codex MRLs for some pesticide residues in some grains.These residues were dispersed among the four classes of pesticides: carbamates, organochlorines, organophosphates, and pyrethroids. In actuality, 90% of the mainly banned organochlorine pesticides exceeded MRLs. Thus, this study revealed concentration levels of organo-chloride pesticides and organo-phosphate pesticides in grain samples drawn from selected markets in southwest Nigeria. Consumers seeking high-quality food in Nigeria should take note of these facts. Notwithstanding, the small percentage of samples with detectable residues suggests that there is a need to increase the monitoring of pesticides in grains, educate farmers, and raise their awareness of the dangers of unauthorized use of pesticides that are restricted for use in agriculture, which can harm the industry’s reputation as a whole.

## 1. Introduction

People who live in rural areas are frequently exposed to pesticides, solvents, and metals, as well as other environmental risks linked to agricultural output. According to reports, chronic exposure to pesticides can cause conditions like diabetes, cancer, cardiovascular disease, and neurological issues [1-11]. Consequently, there is now much greater awareness about the health concerns associated with exposure to pesticides [1, 5,4, 5, 8-10]. With the world population projected to reach about 10 billion people by 2050, increasing food production is every nation’s top priority [12-15]. Evidence suggests that the world’s population is growing by 97 million people yearly. In fact, the Food and Agricultural Organization (FAO) of the United Nations has made the grim prediction that in order to meet the demand of a growing population, global food production must expand by 70% [16-18]. However, increasing food production is beset by a number of increasingly difficult problems, not the least of which is the extremely limited expansion of cultivable land. Because of this, the existing agricultural system is under immense pressure to produce enough food using the same amount of land, water, and other resources as before [19-22]. Herbicides, insecticides, fungicides, nematicides, fertilizers, and soil amendments are currently utilized in greater amounts than in the past in order to increase crop production [1-9]. Since the development of synthetic insecticides in 1940, when organochlorine (OCl) pesticides were first applied for pest control, these chemicals have mostly entered the scene. Prior to this introduction, the majority of weeds, pests, insects, and illnesses were managed utilizing environmentally friendly techniques such cultural, mechanical, and physical control methods [2, 1-9]. Pesticides are now a necessary component of modern life and are used to protect agricultural land, food storage facilities, flower gardens, as well as to get rid of pests that spread infectious diseases that are dangerous to humans [1-9, 23-29]. Pesticides are thought to cost roughly $38 billion worldwide each year [30]. To address the need on a global scale, manufacturers and researchers are developing novel pesticide formulations. The applied pesticides should ideally only be poisonous to the target organisms, be somewhat biodegradable, and environmentally beneficial [22, 31]. Unfortunately, this is rarely the case because the majority of pesticides are general-purpose and may kill organisms that are beneficial to the ecosystem or harmless. The majority of pesticides are thought to poison the environment, with just around 0.1% of them reaching the intended target organisms [1-9, 32]. Numerous elements of the water, air, and soil ecosystem have been contaminated by the continual application of persistent and non-biodegradable pesticides [1-9]. Additionally, pesticides have bioaccumulated in the upper tropic level of the food chain [16, 22]. More recently, exposure to pesticides has been linked to a number of acute and chronic human disorders [1-10, 33]. Many people are well aware that pesticide chemicals frequently come into contact with them through food residues [4]. Since pesticides are frequently utilized due to their capacity to destroy biological species like insects, plant diseases, and weeds, many consumers may find this knowledge to be upsetting. The topic of what risks, if any, consumers are exposed to as a result of consuming pervasive pesticide residues in their diet is raised by their existence in food. Advocates frequently give advice to consumers regarding safety issues, such as whether to eat conventional or organic foods, which geographic sources of foods should be avoided, and what may be done to decrease exposure to pesticides by washing, peeling, boiling, or scrubbing foods before consumption [1-9]. Such recommendations usually lack the necessary scientific support. Since pesticides, like all chemicals, adhere to the rules of toxicology, the mere presence of a pesticide residue on a food item does not prove that it has caused harm. The potential dangers of a chemical are determined by its dose or exposure from series of studies [34-51]. Contrary to the wise counsel from healthcare professionals that calls for balanced and diverse diet rich in whole grains, consumers may be opting to heed dubious advice to stay away from particular foods because of possible pesticide residues. Studies show that residues are frequently found, and that people regularly come into contact with pesticides in their food [2, 3, 6, 7]. It is frequently implied that pesticide residue discoveries that contravene NAFDAC food safety standards may endanger the health of consumers because the relationship between permissible levels of pesticides on foods (tolerances) and safety is poorly understood [4]. Despite having a population of 26 million, Nigeria ranks fourth in the world for the number of undernourished individuals [52]. Due to their high nutritious content, staple foods including maize, rice, millet, and cowpea present a chance to reduce malnutrition. For instance, compared to animal products, cowpeas are a cheap source of protein whereas maize is high in carbohydrates [53]. However, malnutrition has been associated with relying too heavily on one or a few of these basics [54]. In order to optimize mineral bioavailability and fortification, it would be helpful to understand the nutritional makeup of these grains. The environment, animal products, cash crops, vegetables, and processed foods have been the main subjects of the most recent studies on pesticide residues in food in Nigeria. Additionally, research on cereals like maize has only been done in a confined region [55]. A thorough analysis of pesticide residues in common foods in various states is required to determine potential dangers and exposure and to protect the general public’s health in light of the requirement for regular monitoring of toxins in food [56-60]. There are significant concerns about the health risks associated with exposure to many pesticides during employment as well as consumer exposure to their residues in food and drinking water [34-51], despite the fact that the procedures for the approval of active substances of pesticides for agricultural use adhere to strict regulations to ensure both their high effectiveness and their environmental and consumer safety [4]. The irrational use of pesticides, which can have both immediate and long-term repercussions on human health and the environment, is frequently linked to the presence of pesticide residues in agricultural crops [1-9]. Numerous pesticides and fungicides used to control crop diseases and pests are now known to be carcinogenic, harmful to developing fetuses, endocrine disruptors, and to increase the risk of immunological, neurological, and respiratory illnesses [1-9].

Numerous research have confirmed the link between pesticide use and an elevated risk of different malignancies as well as other health issues [1-9]. Young infants are particularly at risk from pesticide exposure connected to various sources, making them susceptible to potential neurological, neurodevelopmental, and other consequences [1-9]. The development of a child’s brain and central nervous system can be hampered by even minor early-life pesticide exposure [9]. Consumers who desire food free of agricultural residues are concerned about the hazards from exposure to pesticides that are all mentioned above and others [2-10]. The majority of consumers believe that grains are safe and good for their health, and they link this belief to the lack of agricultural chemical residues, as well as to a natural flavor and concern for the environment. Its restricted production practices are the cause of these perceived qualities, which increase consumer trust across the globe. In addition, a significant and essentially constant proportion of food products with detectable pesticide residues are present on the European market, according to reports on the level of pesticide contamination in food. The tests conducted yearly as part of the mandated control programs encompassing the entire EU and affiliated nations have brought this issue to light in recent years. The European Food Safety Authority is in charge of this market surveillance throughout Europe (EFSA). According to the EFSA annual reports, between 39.5 and 45.5% of the tested samples had detectable pesticide residues in recent years. Additionally, between 2.6 and 5.1% of the samples included residues that were above the permitted limits (referred to as the MRL-maximum residue level in food of plant origin). Nevertheless, EFSA monitoring shows that a portion of organic samples contain detectable pesticide residues even though the use of synthetic pesticides in organic agriculture is explicitly prohibited by EU Regulation 2018/848. 8.3% of organic samples had detectable pesticide residues in 2015, and 19.9% did so in 2020. Of the analyzed organic samples, 0.7% had residues above the MRL in 2015, and 1.5% did so in 2020. This paper’s objective is to provide and calculate the discovered pesticide’s concentration in grains using maximum residue limits (MRL). Consumers, who are increasingly looking for high-quality, pesticide-free grains, may be quite interested in the findings on the safety status of these popular grains originating from the chosen market in the southwest region of Nigeria. When purchasing grains, consumers have a right to anticipate what they receive will be of good nutritional and biological value and won’t be harmful to their health. Farmers must adhere to national and international laws and regulations in order to safeguard consumers and themselves from a variety of risks. Food safety precautions begin with farm production and continue all the way through the supply chain [4-8].

## 2. Material and Methods

### Study Area

The study area is selected markets across four states (Ekiti, Oyo, Osun and Ondo States) in Southwestern, Nigeria. The markets were: Oja-titun (market) Ile-Ife (16° 18′ N 23° 33′ E), Osun; Alesinloye Market Ibadan, Oyo (7° 26′ N 3° 55′ E; Oja Oba, Ado-Ekiti, Ekiti (7° 37′ N 5° 13′ E); and Oja Oba, Akure Ondo State (7° 15′ N 5° 12′ E) (Fig. 1 below). Hereinafter, the market will be referred to as Ekiti, Oyo, Osun and Ondo, for clarity.

**Fig. 1:**
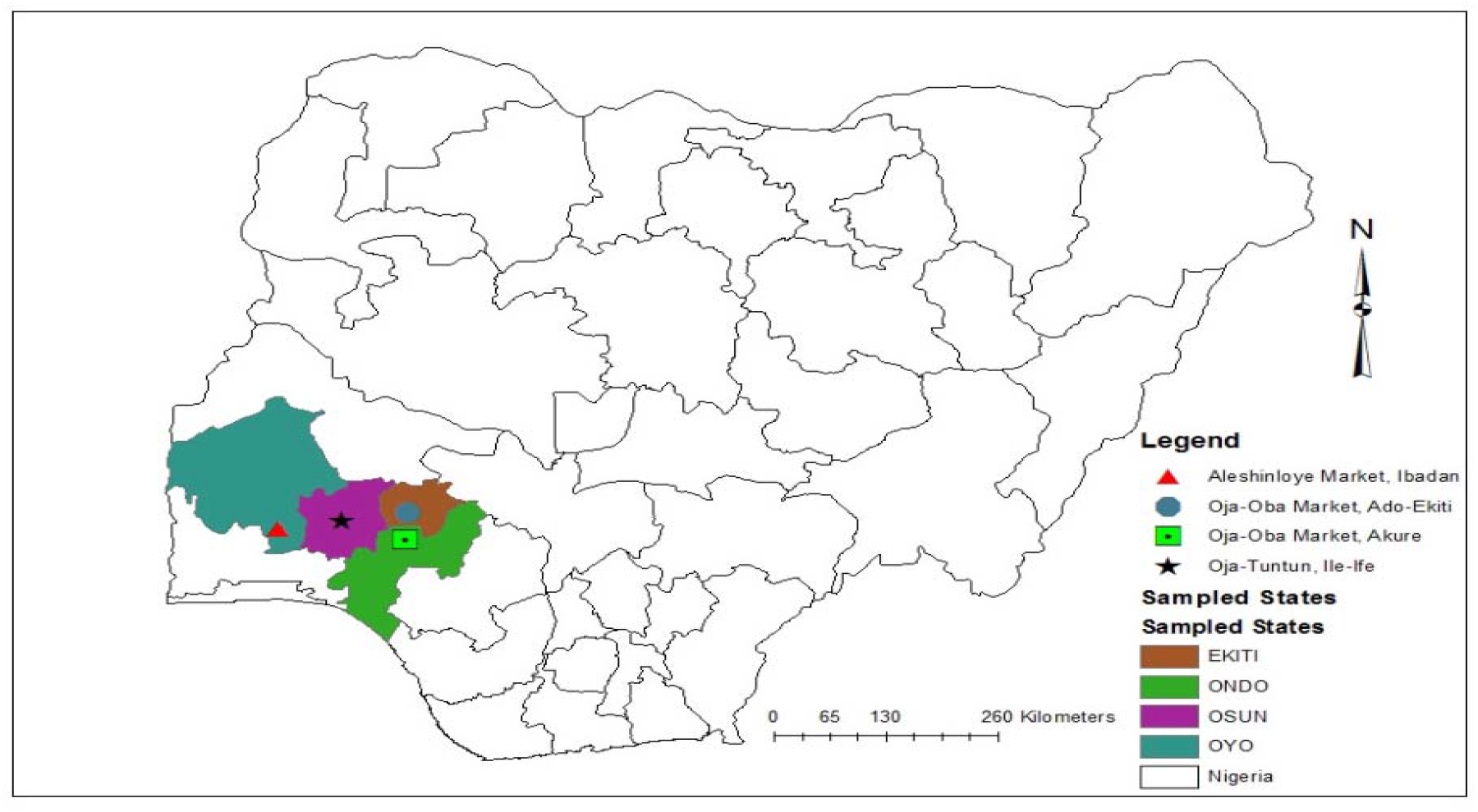
Map showing the study area

### Sample collection and preparation

Samples of dry beans, maize, millet, and rice (local and foreign) were purchased from the four markets. Six samples of: brown and white beans, white and yellow maize, brown millet and rice were collected from all four markets except for Ondo (Oja-Oba market, Akure) where yellow maize was not available. The grain samples were bought in the dried state such that there was no need for the samples to undergo additional drying. We patronized grain merchant and purchased each of the six grains sample. Each grain sample was bought in three separate portions of 200 – 250g and put in black-coloured polyethylene plastic bags, labelled and transported to the laboratory. On arrival, the samples were sorted to remove impurities including stones and shafts. Thereafter, the samples were thoroughly ground using mortar and pestle, and thereafter a hand-grinding machine was used to pulverize into fine powder. Finally, each of the powdered grain sample was stored in labelled Ziploc bag, and kept at 4°C in a refrigerator.

### Proximate analysis

The grain samples were subjected to proximate analysis including ash content, moisture content, crude fat, crude fibre, crude protein, and total carbohydrate [61]. Triplicate samples were constituted by randomly picked samples from three of the four markets. The replicates of each market were pooled.

### Determination of moisture content

Moisture content was determined after drying using the oven. First, the crucibles were cleaned thoroughly and dried in an oven. After cooling, the weights of the crucible were recorded. Thereafter, 1.0g of each sample was weighed and put into the crucible before drying at 105 °C until constant weights were attained. Moisture content was calculated using the formula:

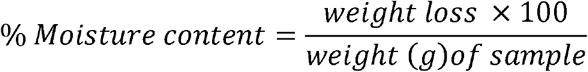

### Determination of ash content

To determine the ash content, each finely ground samples was weighed into a clean, moisture-free crucible. The organic matter was charred by burning over low flame without the lid covering. Thereafter, crucible was transferred to a muffle furnace at 600 °C for 6h. Following this, the samples became completely ashed. It was then cooled in a desiccator and reweighed.

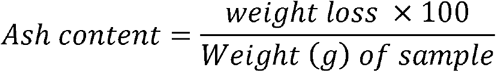

### Determination of crude protein

The micro-Kjeldah method was used in determining the crude protein of the samples [61].

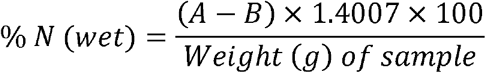

A is the volume (ml) of std HCl × the normality of std HCl B is the volume (ml) of std NaOH × normality of std NaOH

### Determination of crude fat

The crude fat content of each sample was determined using Soxhlet apparatus. 10g of each sample added to a pre-weighed filter paper. The filter paper was folded carefully, moved into a pre-weighed thimble and reweighed before the transfer into the Soxhlet apparatus for extraction using n-hexane at 60 – 80 ° C for 6 hours. Thereafter, the solvent evaporated after exposing to 100 ° C in an oven for half an hour. On cooling (in a desiccator), the thimble was reweighed.

The fat extracted from a given quantity of sample was then calculated:

This was carried out gravimetrically. 5g of the sample was weighed into thimble. Petroleum ether was used for extraction at 60 – 80 °C for 3 hours. The solvent was evaporated and the flask reweighed.

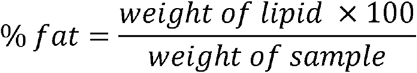

### Determination of Crude fibre

A portion of the ether extract (fatless), was used in crude fibre determination. First, dilute acid was used in the heating the ether extract This was then serially heated with dilute acid and then hydrolyzed with dilute alkali so the digestible portion is negated. Finally, the difference between the weight of the residue and the ash was recorded as the fibre content [61].

### Determination of total carbohydrate

The percentage carbohydrate content of the samples was determined indirectly from other proximate composition as follows:

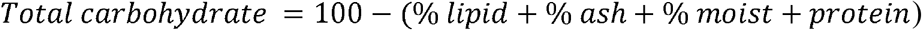

### Determination of pesticide residues Chemicals and reagents

Commercial standards of Quinalphos, Diazinon, Fenitrothion, Dimethoate, Acephate, Malathion and Chlorpyrifos were gotten from Sigma Aldrich Laborchemikalien (St Louis, MO, USA) through Top Scientific Ltd. Ado-Ekiti Ekiti state, Nigeria. They all had >99.6% purity. Other regents including sodium chloride (NaCl), acetone, methanol, anhydrous magnesium sulphate (MgSO_4_), gradient grade acetonitrile and Primary Secondary Amine (PSA) were purchased from Top Scientific Ltd. Ado-Ekiti Ekiti state, Nigeria.

### Preparation of pesticide standard solution

The stock solutions of each of the aforementioned commercial standard reagents were prepared separately in acetone (1000 mg/L). A mixed standard solution (50 mg/L actetone) containing the standard reagents was prepared from the individual stock solution and then a lower concentration of 10 mg/L. Thereafter 0.1, 0.2, 0.5, 1.0, 2.0, 3.0, and 5.0 mg/L working standard were prepared by measuring the required volume to make up each of the concentration in a 10ml flask and making it up with the stock solution of acetone.

### Extraction and clean up

QuEChERS extraction was carried out as described by the European Committee for Standardization [62]. First, 10 g of the homogenized sample was weighted into a 50 mL polypropylene centrifuge tube, 10 mL of acetonitrile (MeCN) was added followed by the use of a vortex mixer to shake the centrifuge for 30s. NaCl (1 g) and Anhydrous MgSO_4_ (4g) were added followed by immediate shaking with the vortex mixer for 60 seconds to pre-empt the MgSO_4_ from becoming aggregates and then, the extract was centrifuged at 5000 rpm for 5 minutes. A 3 mL aliquot of the MeCN layer was moved into a 15 mL micro centrifuge tube holding 600 mg of and 120 mg of anhydrous MgSO_4_ and Primary Secondary Amine, respectively. The vortex was again used to thoroughly 30 s before centrifuged at 4000 rpm for 5 minutes. Finally, a 0.2 µm PTFE filter was applied in filtering 1 mL supernatant before taking in a clean vial for injection.

### Instrumentation and methodology of GC-MS analysis

A Varian 3800/4000 gas chromatograph mass spectrometer was used to analyze pesticide residues. The capillary column was set at AT-1 and a length of 30 m, while the ID and film thickness were set at 0.25 mm 0.25 µm, respectively. The GC initial temperature of the GC column was 70 °C and increase to 300 °C after two minutes holding time. At 300 °C, it was subjected to another 7 minutes holding time. Together, the total run time was 32 minutes. Nitrogen (99.9995% purity) was the carrier gas of choice. The GC-MS interface temperature was at 280 °C and carrier gas had a constant flow rate of 1.51 ml/min. Injector and detector temperatures were set at 250 °C, linear velocity was the flow control mode; and a 1μl of sample was injected in the split ratio of 30:0. The range of the MS was 30 – 800 Da. The concentration of pesticide residues in each sample were obtained following the comparative peak retention times of the samples relative to the pure analytical standards; and were represented as relative area percentage. For the identification, the spectrum obtained through GC – MS compounds were compared with the database of the National Institute Standard and Technology. As a precaution to, samples were randomly and continuously injected as a batch in order to separate technical variations from the biological. More so, the prepared pooled samples were injected into the machine at regular intervals as quality control. The concentrations of the pesticide residues were given in mg/kg.

### Survey

Structured questionnaires were administered to 60 respondents in each of the four markets [63]. The questionnaire included questions relating to the: socio-demographic information, attitudes of grain merchants towards grains preservation, knowledge of grain merchants towards pesticide application and the health implication of pesticide usage on humans Appendix.

### Statistical analyses and calculations

Data analyses were carried out on SPSS version 21.0, IBM, USA. The differences between the treatments were evaluated using Analysis of variance (ANOVA) while Duncan’s new multiple-range test was used to separate means at *P* < 0.05. All graphs were plotted on Microsoft Excel, 2019.

## 3. Results

### Concentration of Identified Pesticide in Grains Organophosphates

The OPP residues in the bean samples were in the range of 0.0.14 – 0.036 mg kg^−1^, 0.014 – 0.042 mg kg^−1^, 0.010 – 0.014 mg kg^−1^, 0.011 – 0.045 mg kg^−1^ and 0.022 – 1.230 mg kg^−1^ for Phenthoate, Chlorthiophos, Ethion, Prothiofos and Iodofenphos, respectively (Table 2 & 3). Compared to the MRL set by the EU, the level of Ethion in the bean samples was lesser or equal to the stipulated value of 0.01. Only brown beans from Ondo and White beans from Ado-Ekiti were equal to this value.

**Table 2:** Mean concentrations (mg kg^−1^) of organochlorine pesticide residues in bean samples obtained from selected markets in Southwest Nigeria.

| Pesticide residue | Concentration ($\text{mg kg}^{-1}$) | | | | | |
| --- | --- | --- | --- | --- | --- | --- |
|  | MRL<sub>1</sub> | MRL<sub>2</sub> | Ado-Ekiti | Ibadan | Osun | Ondo |
| **Brown beans** |  |  |  |  |  |  |
| δ-Lindane | 0.01 | 0.01 | $0.012 \pm 0.00$ | $0.026 \pm 0.00$ | $0.012 \pm 0.00$ | $0.027 \pm 0.00$ |
| α-Lindane | 0.01 | 0.01 | $0.032 \pm 0.00$ | $0.032 \pm 0.00$ | $0.027 \pm 0.00$ | $0.033 \pm 0.00$ |
| β-Hexachlorocyclohexane | 0.01 | 0.01 | $0.013 \pm 0.00$ | $0.010 \pm 0.00$ | $0.026 \pm 0.00$ | $0.012 \pm 0.00$ |
| p,p'-DDT | 0.1 | 0.05 | $0.200 \pm 0.01$ | $0.200 \pm 0.02$ | $0.230 \pm 0.00$ | $0.249 \pm 0.01$ |
| Dieldrin | 0.02 | 0.01 | $0.288 \pm 0.00$ | $0.024 \pm 0.00$ | $0.100 \pm 0.08$ | $0.022 \pm 0.00$ |
| p,p'-DDE | 0.1 | 0.05 | $0.190 \pm 0.00$ | $0.209 \pm 0.00$ | $0.173 \pm 0.00$ | $0.187 \pm 0.01$ |
| Aldrin | 0.02 | 0.01 | $0.012 \pm 0.00$ | $0.011 \pm 0.00$ | $0.010 \pm 0.00$ | $0.013 \pm 0.00$ |
| **White beans** |  |  |  |  |  |  |
| δ-Lindane | 0.01 | 0.01 | $0.022 \pm 0.00$ | $0.022 \pm 0.00$ | $0.035 \pm 0.00$ | $0.031 \pm 0.00$ |
| α-Lindane | 0.01 | 0.01 | $0.036 \pm 0.00$ | $0.030 \pm 0.00$ | $0.034 \pm 0.00$ | $0.033 \pm 0.00$ |
| β-Hexachlorocyclohexane | 0.01 | 0.01 | $0.043 \pm 0.00$ | $0.031 \pm 0.00$ | $0.036 \pm 0.00$ | $0.023 \pm 0.00$ |
| p,p'-DDT | 0.1 | 0.05 | $0.217 \pm 0.00$ | $0.211 \pm 0.00$ | $0.213 \pm 0.00$ | $0.214 \pm 0.00$ |
| Dieldrin | 0.02 | 0.01 | $0.253 \pm 0.00$ | $0.253 \pm 0.00$ | $0.016 \pm 0.00$ | $0.026 \pm 0.00$ |
| p,p'-DDE | 0.1 | 0.05 | $0.196 \pm 0.00$ | $0.213 \pm 0.01$ | $0.349 \pm 0.00$ | $0.210 \pm 0.01$ |
| Aldrin | 0.02 | 0.01 | $0.012 \pm 0.00$ | $0.013 \pm 0.00$ | $0.017 \pm 0.00$ | $0.011 \pm 0.00$ |

MRL<sub>1</sub> and MRL<sub>2</sub> are the maximum residue limit ( $\text{mg kg}^{-1}$ ) set by the FAO/WHO and the European Union, respectively. Values are expressed as mean ± S.E. (n=3).

**Table 3:** Mean concentrations (mg kg^−1^) of organophosphate pesticide residues in beans samples obtained from selected markets in Southwest Nigeria

| Pesticide residue | Concentration ($\text{mg kg}^{-1}$) | | | | | |
| --- | --- | --- | --- | --- | --- | --- |
|  | MRL<sub>1</sub> | MRL<sub>2</sub> | Ado-Ekiti | Ibadan | Osun | Ondo |
| **Brown beans** |  |  |  |  |  |  |
| Phenthoate | N/A | N/A | $0.023 \pm 0.00$ | $0.034 \pm 0.00$ | $0.014 \pm 0.00$ | $0.033 \pm 0.00$ |
| Chlorthiophos | N/A | N/A | $0.024 \pm 0.00$ | $0.022 \pm 0.00$ | $0.020 \pm 0.00$ | $0.019 \pm 0.00$ |
| Ethion | N/A | 0.01 | $0.014 \pm 0.00$ | $0.012 \pm 0.00$ | $0.012 \pm 0.00$ | $0.010 \pm 0.00$ |
| Prothiofos | N/A | N/A | $0.016 \pm 0.01$ | $0.013 \pm 0.00$ | $0.017 \pm 0.00$ | $0.012 \pm 0.00$ |
| Iodofenphos | N/A | N/A | $0.022 \pm 0.00$ | $0.024 \pm 0.00$ | $0.022 \pm 0.00$ | $0.022 \pm 0.00$ |
| **White beans** |  |  |  |  |  |  |
| Phenthoate | N/A | N/A | $0.030 \pm 0.00$ | $0.025 \pm 0.00$ | $0.036 \pm 0.00$ | $0.025 \pm 0.00$ |
| Chlorthiophos | N/A | N/A | $0.024 \pm 0.00$ | $0.021 \pm 0.00$ | $0.042 \pm 0.00$ | $0.014 \pm 0.00$ |
| Ethion | N/A | 0.01 | $0.010 \pm 0.00$ | $0.013 \pm 0.00$ | $0.011 \pm 0.00$ | $0.011 \pm 0.00$ |
| Prothiofos | N/A | N/A | $0.023 \pm 0.00$ | $0.045 \pm 0.00$ | $0.035 \pm 0.00$ | $0.011 \pm 0.00$ |
| Iodofenphos | N/A | N/A | $0.022 \pm 0.00$ | $0.023 \pm 0.00$ | $1.230 \pm 0.01$ | $0.027 \pm 0.00$ |

MRL<sub>1</sub> and MRL<sub>2</sub> are the maximum residue limit ( $\text{mg kg}^{-1}$ ) set by the FAO/WHO and the European Union, respectively. “N/A” means not available. Values are expressed as mean ± S.E. (n=3).

### Pyrethroids

The concentration of Amitraz and Cypermethrin I were lower that the EU’s MRL in both brown and white beans (Table 4). Compared to EU’s MRL of 0.02 mg kg^−1^, white beans had higher levels of Flumioxazin across all markets except for Osun while Ado-Ekiti and Osun were the only ones that exceeded the EU MRL in the case of brown beans.

**Table 4:** Mean concentrations (mg kg^−1^) of pyrethroid and carbamate pesticide residues in bean samples obtained from selected markets in Southwest Nigeria

| Pesticide group | Pesticide residue | Concentration (mg kg<sup>-1</sup>) |  |  |  |  |  |
| --- | --- | --- | --- | --- | --- | --- | --- |
|  |  | MRL<sub>1</sub> | MRL<sub>2</sub> | Ado-Ekiti | Ibadan | Osun | Ondo |
| Brown beans |  |  |  |  |  |  |  |
| Pyrethroid | Amitraz | N/A | 0.05 | 0.016 ± 0.00 | 0.014 ± 0.00 | 0.012 ± 0.00 | 0.016 ±0.00 |
|  | Flumioxazin | N/A | 0.02 | 0.021 ± 0.00 | 0.020 ± 0.00 | 0.026 ± 0.00 | 0.016 ± 0.00 |
|  | Cypermethrin I | N/A | 0.7 | 0.012 ± 0.00 | 0.015 ± 0.00 | 0.015 ± 0.00 | 0.014 ± 0.00 |
| Carbamate | Carbofuran | N/A | 0.01 | 0.003 ± 0.00 | 0.019 ± 0.00 | 0.006 ± 0.00 | 0.013 ± 0.00 |
| White beans |  |  |  |  |  |  |  |
| Pyrethroid | Amitraz | N/A | 0.05 | 0.011 ± 0.00 | 0.014 ± 0.00 | 0.021 ± 0.00 | 0.015 ± 0.00 |
|  | Flumioxazin | N/A | 0.02 | 0.021 ± 0.00 | 0.022 ± 0.00 | 0.017 ± 0.00 | 0.031 ± 0.00 |
|  | Cypermethrin I | N/A | 0.7 | 0.010 ± 0.00 | 0.014 ± 0.00 | 0.015 ± 0.00 | 0.015 ± 0.00 |
| Carbamate | Carbofuran | N/A | 0.01 | 0.010 ± 0.00 | 0.005 ± 0.00 | 0.025 ± 0.00 | 0.020 ± 0.00 |

MRL<sub>1</sub> and MRL<sub>2</sub> are the maximum residue limit (mg kg<sup>-1</sup>) set by the FAO/WHO and the European Union, respectively. “N/A” means not available. Values are expressed as mean ± S.E. (n=3).

### Carbamates

Carbofuran was the only carbamate found in beans samples (Table 4). Its concentration was higher the MRL stipulated by the EU in beans samples collected from Ondo as well as the brown beans from Ibadan and white beans from Osun.

### Maize Organochlorine

Table 5 shows the mean concentrations of OCP residues in maize samples. δ-Lindane and β-Hexachlorocyclohexane were higher than the MRL of 0.01 mg kg^−1^ set by the FAO/WHO and the EU in both yellow and white maize in all the markets. Likewise, the level of p,p’-DDT, Dieldrin were also higher than the MRL in white maize across all the markets and yellow maize in Ado-Ekiti, Ibadan and Ondo. While the concentrations of dieldrin in white maize of all the markets was higher than the MRL set by both the FAO/WHO and the EU, its concentration in yellow maize was lower than the FAO/WHO MRL but higher than the EU standard. On the bright side, the concentrations of Aldrin and Endosulfan in the maize samples across all the markets were lower than the MRL of 0.05 mg kg^−1^ set by the EU except for yellow maize samples collected from Ondo.

**Table 5:** Mean concentrations (mg kg^−1^) of organochlorine pesticide residues in maize samples obtained from selected markets in Southwest Nigeria

| Pesticide residue | Concentration (mg kg<sup>-1</sup>) |  |  |  |  |  |
| --- | --- | --- | --- | --- | --- | --- |
| Pesticide residue | Concentration (mg kg<sup>-1</sup>) |  |  |  |  |  |
|  | MRL<sub>1</sub> | MRL<sub>2</sub> | Ado-Ekiti | Ibadan | Osun | Ondo |
| **Yellow maize** |  |  |  |  |  |  |
| δ-Lindane | 0.01 | 0.01 | 0.025 ± 0.00 | 0.025 ± 0.00 | 0.123 ± 0.00 | 0.021 ± 0.00 |
| β-Hexachlorocyclohexane | 0.01 | 0.01 | 1.004 ± 0.00 | 1.000 ± 0.00 | 1.003 ± 0.00 | 0.999 ± 0.00 |
| p,p'-DDT | 0.1 | 0.05 | 2.170 ± 0.02 | 2.157 ± 0.00 | 0.064 ± 0.00 | 2.157 ± 0.00 |
| Dieldrin | 0.02 | 0.01 | 0.013 ± 0.00 | 0.013 ± 0.00 | 0.014 ± 0.00 | 0.014 ± 0.00 |
| Aldrin | 0.02 | 0.01 | 0.011 ± 0.00 | 0.012 ± 0.00 | 0.012 ± 0.00 | 0.017 ± 0.00 |
| Endosulfan | N/A | 0.05 | 0.016 ± 0.00 | 0.015 ± 0.00 | 0.018 ± 0.00 | 0.070 ± 0.05 |
| **White maize** |  |  |  |  |  |  |
| δ-Lindane | 0.01 | 0.01 | 0.104 ± 0.00 | 0.026 ± 0.00 | 0.982 ± 0.00 |  |
| β-Hexachlorocyclohexane | 0.01 | 0.01 | 1.000 ± 0.00 | 0.998 ± 0.00 | 0.026 ± 0.00 |  |
| p,p'-DDT | 0.1 | 0.05 | 0.017 ± 0.00 | 0.027 ± 0.00 | 0.019 ± 0.00 |  |
| Dieldrin | 0.02 | 0.01 | 0.029 ± 0.00 | 1.111 ± 0.00 | 0.021 ± 0.00 |  |
| Aldrin | 0.02 | 0.01 | 0.012 ± 0.00 | 0.016 ± 0.00 | 0.010 ± 0.00 |  |
| Endosulfan | N/A | 0.05 | 0.022 ± 0.00 | 0.012 ± 0.00 | 0.013 ± 0.00 |  |

MRL<sub>1</sub> and MRL<sub>2</sub> are the maximum residue limit (mg kg<sup>-1</sup>) set by the FAO/WHO and the European Union, respectively. “N/A” means not available. Values are expressed as mean ± S.E. (n=3). White maize was unavailable in Ondo market at the time of sample collection.

### Organophosphates

The mean concentrations of the OPP residues are presented in Table 6. The concentration of Chlorthiophos, which was detected only in yellow maize, was between 0.018 and 0.049 mg kg^−1^. The minimum and maximum values were observed in Osun and Ado-Ekiti, respectively. Similarly, Ethion was not detected in all white and yellow maize samples as well as in yellow maize collected from Osun. Its concentrations in samples from Ondo were lower was higher than the EU’s MRL in samples obtained from Ondo. The levels of Malathion were higher than the FAO/WHO recommended MRL of 0.05 mg kg^−1^ in all samples of yellow and white maize obtained from Ado-Ekiti and Ibadan. The concentrations of Dichlorvos in both yellow and white maize samples were higher with MRL of 0.01 mg kg^−1^ set by the EU.

**Table 6:**
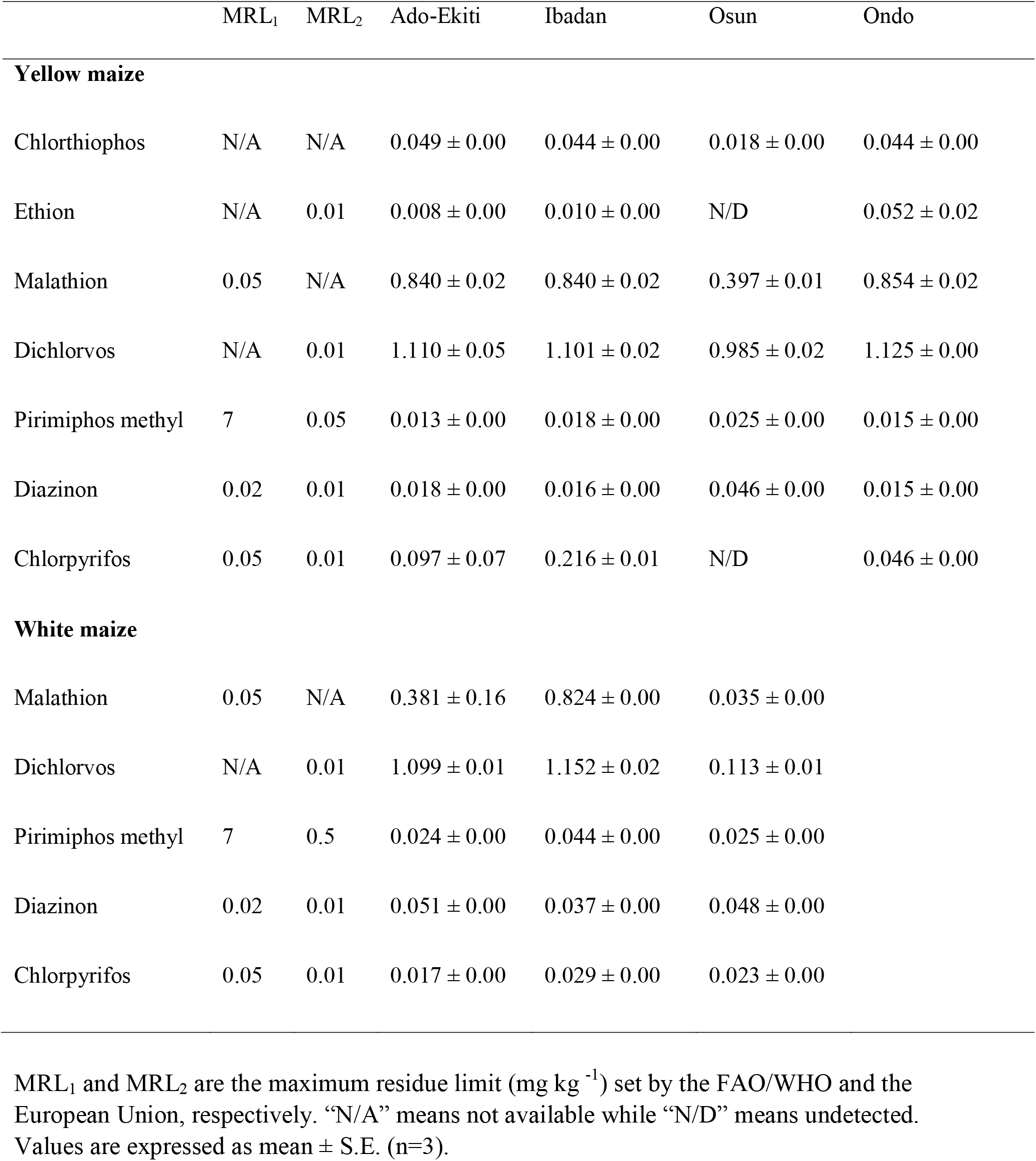
Mean concentrations (mg kg^−1^) of organophosphate pesticide residues in maize samples obtained from selected markets in Southwest Nigeria

Conversely, the levels of pirimiphos methyl in both yellow and white maize samples were well below the MRL stipulated by the FAO/WHO and slightly below that of the EU. Diazinon level in white maize were all higher than the MRLs while those of the yellow maize were all higher than the EU’s MRL but generally lower than that of the FAO/WHO. Diazinon levels in maize collected from Osun had a mean value of 0.046 mg kg^−1^ which was very much higher that the stipulated MRLs. Chlorpyrifos was highest in yellow maize obtained from Ibadan but was not detected in Osun. In addition to Ibadan, its concentration in yellow maize from Ado-Ekiti was higher than MRLs. On the other hand, its concentrations in white maize across all markets were lower than the MRL set by FAO/WHO (0.05 mg kg^−1^) but higher that of the EU (0.01 mg kg^−1^).

### Pyrethroids

Amitraz was the only pyrethroid found in the maize sample (Table 7). Its concentration, which was in the range of 0.003 – 0.034 mg kg^−1^, was lower than the EU MRL of 0.05 mg kg^−1^.

**Table 7:**
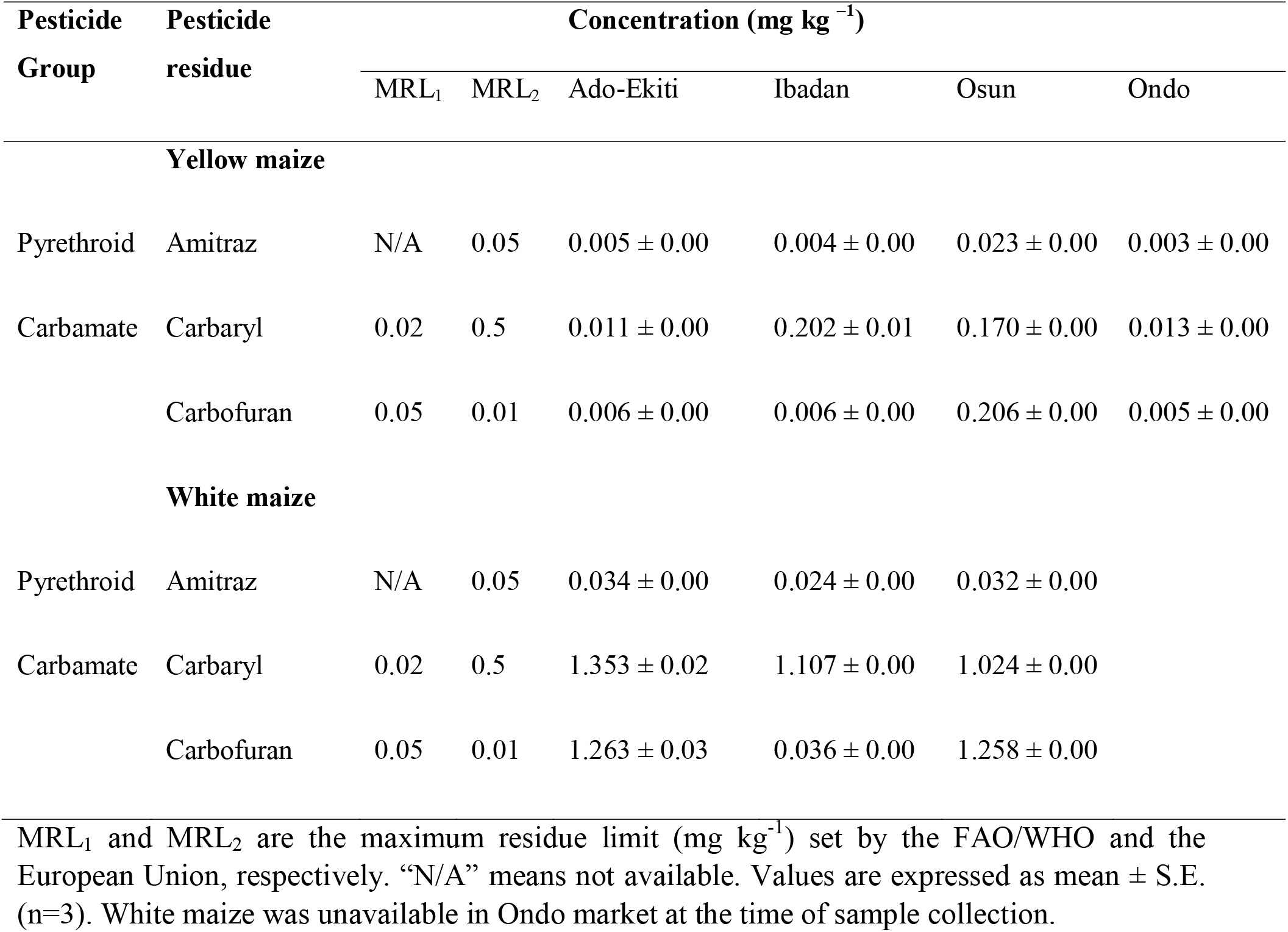
Mean concentrations (mg kg^−1^) of pyrethroid and carbamate pesticide residues in maize samples obtained from selected markets in Southwest Nigeria

### Carbamates

The concentration of Carbaryl in yellow maize obtained from Ibadan and Osun were higher than the MRLs (Table 7). In the case of white maize, all samples across the markets recorded a range of 1.024 – 1.353 mg kg^−1^, which were significantly higher than the 0.05 mg kg^−1^ and 0.02 mg kg^−1^ MRLs set by the FAO/WHO and EU, respectively. The concentration of Carbofuran in yellow maize for all the selected markets were lower than the MRLs. However, only Osun had a higher concentration of Carbofuran (0.206 mg kg^−1^) when compared with FAO/WHO MRL of 0.01 mg kg^−1^ or EU’s MRL of 0.05 mg kg^−1^. On the other hand, white maize had a concentration of Carbofuran that was higher than the MRLs across all the four markets.

### Millet Organochlorine

The mean concentration of OCP residues in millet is presented in Table 8. More often than not, the levels of OCP residues in millet grains were higher that the MRL recommended by both the FAO/WHO as well as the EU. First, δ-Lindane, α-Lindane, β-Hexachlorocyclohexane and p,p’-DDT where higher than the MRLs. As for p,p’-DDE, Ado-Ekiti, Ibadan and Ondo all exceeded this limits substantially. While Dieldrin and Heptachlor levels in all millet samples (0.040 – 0.127 mg kg^−1^ and 0.020 – 0.046 mg kg^−1^, respectively) were all higher than EU’s MRL of 0.01 mg kg^−1^, none was over the 0.2 mg kg^−1^ set by the FAO/WHO. Aldrin followed a similar pattern but the 0.021 mg kg^−1^ concentration of millet obtained from Osun was higher than both MRLs. Chlordan was between the range of 0.007 and 0.035 mg kg^−1^.

**Table 8:** Mean concentrations (mg kg^−1^) of organochlorine pesticide residues in millet samplesobtained from selected markets in Southwest Nigeria

| Pesticide residue | Concentration (mg kg<sup>-1</sup>) |  |  |  |  |  |
| --- | --- | --- | --- | --- | --- | --- |
|  | MRL<sub>1</sub> | MRL<sub>2</sub> | Ado-Ekiti | Ibadan | Osun | Ondo |
| δ-Lindane | 0.01 | 0.01 | 0.015 ± 0.00 | 0.016 ± 0.00 | 0.011 ± 0.00 | 0.018 ± 0.00 |
| α-Lindane | 0.01 | 0.01 | 0.044 ± 0.00 | 0.044 ± 0.00 | 0.045 ± 0.00 | 0.146 ± 0.00 |
| β-Hexachlorocyclohexane | 0.01 | 0.01 | 0.025 ± 0.00 | 0.031 ± 0.00 | 0.032 ± 0.00 | 0.036 ± 0.00 |
| p,p'-DDT | 0.1 | 0.05 | 0.103 ± 0.00 | 0.103 ± 0.00 | 0.180 ± 0.01 | 0.037 ± 0.00 |
| Dieldrin | 0.2 | 0.01 | 0.040 ± 0.00 | 0.042 ± 0.00 | 0.046 ± 0.00 | 0.127 ± 0.00 |
| p,p'-DDE | 0.1 | 0.05 | 2.123 ± 0.01 | 2.107 ± 0.00 | 0.049 ± 0.00 | 1.127 ± 0.00 |
| Heptachlor | 0.2 | 0.01 | 0.046 ± 0.00 | 0.020 ± 0.00 | 0.036 ± 0.00 | 0.040 ± 0.00 |
| Chlordan | N/A | N/A | 0.007 ± 0.00 | 0.012 ± 0.00 | 0.009 ± 0.00 | 0.035 ± 0.03 |
| Aldrin | 0.2 | 0.01 | 0.012 ± 0.00 | 0.011 ± 0.00 | 0.021 ± 0.00 | 0.012 ± 0.00 |

MRL<sub>1</sub> and MRL<sub>2</sub> are the maximum residue limit (mg kg<sup>-1</sup>) set by the FAO/WHO and the European Union, respectively. “N/A” means not available. Values are expressed as mean ± S.E. (n=3).

### Organophosphates

The concentration of Phenthoate was in the range of 0.035 – 0.046 mg kg^−1^ while Chlorthiophos and Prothiofos were 0.022 – 0.035 mg kg^−1^, and 0.017 – 0.024 mg kg^−1^ (Table 9). The levels of Ethion in the millet grains from all the markets were above the MRL of 0.01 mg kg^−1^ set by the EU.

**Table 9:**
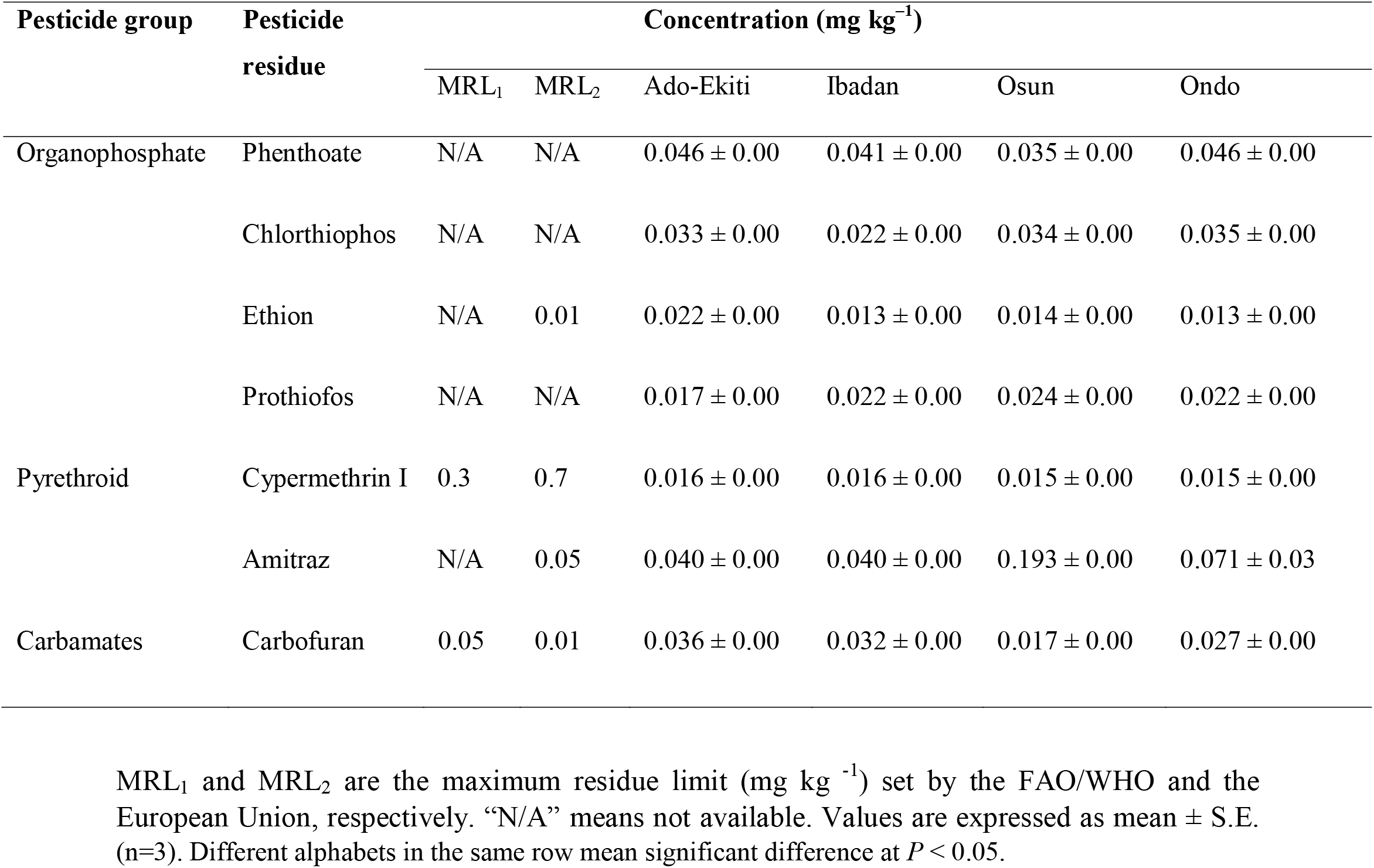
Mean concentrations (mg kg^−1^) of organophosphate, pyrethroid and carbamate pesticide residues in millet samples obtained from selected markets in Southwest Nigeria

| Pesticide group | Pesticide residue | Concentration (mg kg<sup>-1</sup>) |  |  |  |  |  |
| --- | --- | --- | --- | --- | --- | --- | --- |
|  |  | MRL<sub>1</sub> | MRL<sub>2</sub> | Ado-Ekiti | Ibadan | Osun | Ondo |
| Organophosphate | Phenthoate | N/A | N/A | 0.046 ± 0.00 | 0.041 ± 0.00 | 0.035 ± 0.00 | 0.046 ± 0.00 |
|  | Chlorthiophos | N/A | N/A | 0.033 ± 0.00 | 0.022 ± 0.00 | 0.034 ± 0.00 | 0.035 ± 0.00 |
|  | Ethion | N/A | 0.01 | 0.022 ± 0.00 | 0.013 ± 0.00 | 0.014 ± 0.00 | 0.013 ± 0.00 |
|  | Prothiofos | N/A | N/A | 0.017 ± 0.00 | 0.022 ± 0.00 | 0.024 ± 0.00 | 0.022 ± 0.00 |
| Pyrethroid | Cypermethrin I | 0.3 | 0.7 | 0.016 ± 0.00 | 0.016 ± 0.00 | 0.015 ± 0.00 | 0.015 ± 0.00 |
|  | Amitraz | N/A | 0.05 | 0.040 ± 0.00 | 0.040 ± 0.00 | 0.193 ± 0.00 | 0.071 ± 0.03 |
| Carbamates | Carbofuran | 0.05 | 0.01 | 0.036 ± 0.00 | 0.032 ± 0.00 | 0.017 ± 0.00 | 0.027 ± 0.00 |

MRL<sub>1</sub> and MRL<sub>2</sub> are the maximum residue limit (mg kg<sup>-1</sup>) set by the FAO/WHO and the European Union, respectively. “N/A” means not available. Values are expressed as mean ± S.E. (n=3). Different alphabets in the same row mean significant difference at $P < 0.05$ .

### Pyrethroids

The concentration of Cypermethrin I in the millet samples were close and lower than the MRLs set by the FAO/WHO and the EU. On the other hand, millet samples obtained from Ado-Ekiti and Ibadan, which had a common mean concentration of Amitraz (0.040 mg kg^−1^), were lower than the MRL of 0.05 mg kg^−1^ set by the EU while those of Osun and Ondo exceeded this limit.

#### Carbamates

The concentration of Carbofuran, the only identified carbamate in the millet grains, was higher than the MRL of 0.01 set by the EU and lower than the 0.05 mg kg^−1^ set by FAO/WHO across all four markets (Table 9).

#### Rice Organochlorine

The concentration of OCP residues in rice samples is given in (Table 10). δ-Lindane, α-Lindane, exceeded the MRL of 0.01 mg kg^−1^ set by both the FAO/WHO and the EU. β-Hexachlorocyclohexane showed a similar trend as rice samples from all market exceeded the MRL except for rice grains collected in Osun. Meanwhile, the concentration of p,p’-DDT and p,p’-DDE in the rice grains were lower than the MRLs in all the markets. Lastly, Aldrin level in rice grains obtained from Ibadan and Osun were lower than the MRL of 0.01 mg kg^−1^ set by the EU while those of Ondo and Ado-Ekiti higher than this limit but lower than the 0.02 mg kg^−1^ set by the FAO/WHO. The concentration of Dieldrin in any one of the markets was higher than at least one of the MRLs.

**Table 10:** Mean concentrations (mg kg^−1^) of organochlorine pesticide residues in rice samples obtained from selected markets in Southwest Nigeria

| Pesticide residue | Concentration (mg kg<sup>-1</sup>) |  |  |  |  |  |
| --- | --- | --- | --- | --- | --- | --- |
|  | MRL<sub>1</sub> | MRL<sub>2</sub> | Ado-Ekiti | Ibadan | Osun | Ondo |
| δ-Lindane | 0.01 | 0.01 | 0.033 ± 0.00 | 0.035 ± 0.00 | 0.040 ± 0.00 | 0.035 ± 0.00 |
| α-Lindane | 0.01 | 0.01 | 1.031 ± 0.00 | 1.032 ± 0.00 | 1.010 ± 0.00 | 0.699 ± 0.33 |
| β-Hexachlorocyclohexane | 0.01 | 0.01 | 0.018 ± 0.00 | 0.019 ± 0.00 | 0.006 ± 0.00 | 0.020 ± 0.00 |
| p,p'-DDT | 0.1 | 0.05 | 0.010 ± 0.00 | 0.016 ± 0.00 | 0.015 ± 0.00 | 0.011 ± 0.00 |
| Dieldrin | 0.02 | 0.01 | 0.012 ± 0.00 | 0.025 ± 0.00 | 2.112 ± 0.00 | 0.015 ± 0.00 |
| p,p'-DDE | 0.1 | 0.05 | 0.021 ± 0.00 | 0.016 ± 0.00 | 0.013 ± 0.00 | 0.014 ± 0.00 |
| Aldrin | 0.02 | 0.01 | 0.017 ± 0.01 | 0.006 ± 0.00 | 0.004 ± 0.00 | 0.013 ± 0.00 |

MRL<sub>1</sub> and MRL<sub>2</sub> are the maximum residue limit (mg kg<sup>-1</sup>) set by the FAO/WHO and the European Union, respectively. Values are expressed as mean ± S.E. (n=3).

### Organophosphates

Phenthoate, Chlorthiophos, Ethion, Iodophenos and Prothiofos concentrations in the rice grains were in the range of 0.023 – 0.045 mg kg^−1^, 0.005 – 0.026 mg kg^−1^, 1.13 – 1.460 mg kg^−1^, 0.005 – 0.007 mg kg^−1^, 1.50 – 1.528 mg kg^−1^, respectively (Table 11). The minimum mean level of Ethion was at least 100 times higher than the MRL of 0.01 mg kg^−1^ set by the EU.

**Table 11:**
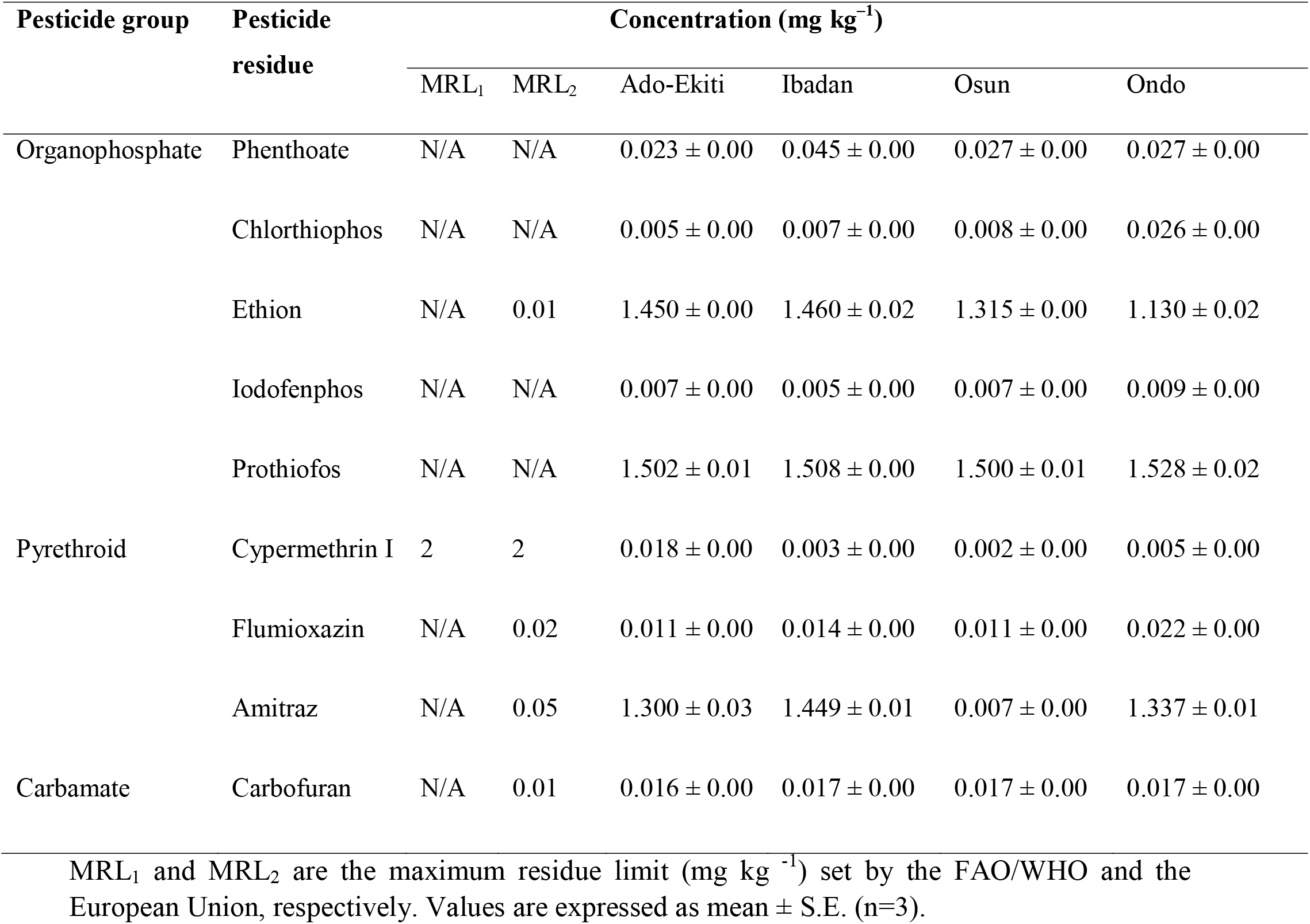
Mean concentrations (mg kg^−1^) of organophosphate, pyrethroid and carbamate pesticide residues in rice samples obtained from selected markets in Southwest Nigeria

#### Pyrethroids

The concentrations of Cypermethrin I in rice was lower than the MRL recommended by the EU (Table 11). Also, Flumioxazin was lower than the set MRL of 0.02 mg kg^−1^ by the EU, except for the rice samples obtained from Ondo. However, Amitraz was generally higher than the set limit of 0.05 mg kg^−1^.

### Carbamates

Only carbofuran was detected in the rice grain samples and its concentration was above the MRL of 0.01 mg kg^−1^ across all markets (Table 11).

## 4. Discussion

### Determine the Concentration of identified pesticides in grains

The worldwide environmental movement began with Rachel Carson, a marine biologist and writer, encouraging the care of nature and preserving the environment, above all, warning about the control of pesticides, so harmful to humans and the environment [64]. In this context, the focus was on the use of DDT (dichlorodiphenyltrichloroethane), an organochlorine insecticide that became important in World War II to combat typhus, yellow fever and malaria-transmitting insects [65]. Its effectiveness in controlling malaria-transmitting insects was so high that between 1946 and 1970 all control programs employed DDT to eradicate the disease. However, due to its high toxicity, there was a decline in the consumption of this insecticide in later years. The Stockholm Convention on Persistent Organic Pollutants emphasizes DDT’s environmental persistence and bioaccumulation, highlighting the search for alternatives in the use of this pesticide [66, 67]. During the last years, it is observed that pesticide residues have spread throughout the environment, contaminating different ecosystems, and compromising food and water resources. It is noted that this contamination comes from population growth, since such growth would not be possible without an increase in food production, and this is closely linked to the use of pesticides and fertilizers [1-9, 68]. Pesticides are part of a large group of organic compounds with different physicochemical characteristics, designed to control and prevent pests in various crops and plantations, improving productivity and is having increasingly strong global impacts on both the environment and human health, often driven by agro-chemicals changes. Numerous reports have emphasized the need for major changes in the global food system: agriculture must meet the twin challenge of feeding a growing population, with rising demand for pesticides, while simultaneously increasing its global environmental impacts. Although, pesticides were used initially to benefit human life through increase in agricultural productivity and by controlling infectious disease, their adverse effects have overweighed the benefits associated with their use. The persistent nature of pesticides has impacted our ecosystem to such an extent that pesticides have entered into various food chains and into the higher trophic levels such as that of humans and other large mammals [1, 11, 16]. Some of the acute and chronic human illnesses have now emerged as a consequence of intake of polluted water [34-51], air [69-74] or food [56-60]. Many of the pesticides have been associated with health and environmental issues [1-10], and the agricultural use of certain pesticides has been abandoned. Exposure to pesticides can be through contact with the skin, ingestion, or inhalation. The type of pesticide, the duration and route of exposure, and the individual health status (e.g., nutritional deficiencies and healthy/damaged skin) are determining factors in the possible health outcome. Within a human or animal body, pesticides may be metabolized, excreted, stored, or bioaccumulated in body fat. The numerous negative health effects that have been associated with chemical pesticides include, among other effects, dermatological, gastrointestinal, neurological, carcinogenic, respiratory, reproductive, and endocrine effects [1-10]. Furthermore, high occupational, accidental, or intentional exposure to pesticides can result in hospitalization and death [2-9]. Residues of pesticides can be found in a great variety of everyday foods and beverages, including for instance cooked meals, water, wine, fruit juices, refreshments, and animal feeds. Furthermore, it should be noted that washing and peeling cannot completely remove the residues. In the majority of cases, the concentrations do not exceed the legislatively determined safe levels. However, these “safe limits” may underestimate the real health risk as in the case of simultaneous exposure to two or more chemical substances, which occurs in real-life conditions and may have synergistic effects. Thus, this study revealed concentration levels of organo-chloride pesticides and organo-phosphate pesticides in grain samples drawn from selected markets in southwest Nigeria. The nutrient composition of cereals is an important indicator of its quality and the overall health benefits of its consumption. First, we observed large variations in moisture content of the different grain samples. These differences can be linked to factors such as planting methods and management practices, processing, environmental factors, pests and diseases etc. [75, 76]. The ash content of grains depends very much on their mineral composition [77]. The ash content of legumes has been generally higher than those of cereals [78, 79]. Similarly, higher ash content in beans was higher than those of the cereals in our study. The protein composition was highest in cowpea and it fell within the range of 20 – 30% reported by Odogwu *et al*. [63]. Also, crude was around 11% reported by Affrifah *et al*. [80]. This is because the removal of the bran and germ of cereals leaves it with the endosperm which lacks nutrients but highly rich in carbohydrate [81]. Pigmented corn was recommended by Libron *et al*. [82] due to it high nutritional value and presence of antioxidants, hence its high protein composition, as against white maize, as observed in our study. However, the higher composition of ash content, crude fibre and crude fat in white maize we observed may be attributed to varietal factors and performance [82]. It is well known that rice has a higher composition of carbohydrate due to its high starch content while maize and millet have better crude protein and fat [83]. In our study as rice also had the highest composition of carbohydrate and which ranged between 76.92 and 86.03% similar to that reported by Oko and Ugwu [84] who compared several varieties of rice in Nigeria. Meanwhile, maize and millet had a higher crude protein and fat levels. While, pesticides are commonly applied in preserving food products in Nigeria. Hence, they have been widely reported in food crops [1-9, 55, 72]. Some OCPs in this study such as β-HCH, Heptachlor, Endosulfan, Aldrin, Lindane, Dieldrin, p,p’-DDE, p,p’-DDT have been previously reported in maize, beans, millet and rice in Nigeria [85-88]. Similarly, Akinneye *et al*. [89] reported Dichlorvos, Carbofuran, Carbaryl, Diazinon, Pirimiphos-methyl, Malathion, Chlorpyrifos as the OPPs and carbamates in beans, rice, wheat and maize samples collected from Ondo State. Additional pesticide residues such as those of Phenthoate and Iodofenphos found in our study could be as a result of the various locations and larger sample size in our work. Amitraz, Cypermethrin and Flumioxazin, which were the pyrethroids residues observed in our study had been reported in grain samples in Nigeria [90]. This suggests the widespread use of various groups of pesticides including organochlorine, organophosphate, carbamates and pyrethroids for grain preservation across Nigeria. In cowpea and maize grains obtained directly from farms in Ghana, Akoto *et al*. [91] reported the mean concentration of organochlorine, organophosphates, and pyrethroids ranging from 0.001 to 0.103 mg kg^−1^, 0.002 to 0.019 mg kg^−1^ and 0.002 to 0.028 mg kg^−1^ in maize, and 0.001 to 0.108 mg kg^−1^, 0.002 to 0.015 mg kg^−1^, 0.001 to 0.039 mg kg^−1^ in cowpea, respectively. Our values were generally higher than the upper limits of these range of values because higher doses of pesticides must have been applied during postharvest storage. The concentration of OCP residues in maize sample in this study were comparable to those of Anzene *et al*. [85] in Nassarawa State which reported a range of 0.03 to 0.13 mg/kg, 0.018 to 0.02 mg/kg, 0.005 to 0.02 mg/kg, 0.25 to 1.25 mg/kg, and 0.04 mg/kg for Aldrin, Dieldrin, Endosulfan, Lindane, and DDT, in maize and guinea corn respectively. While Aldrin and Dieldrin was within these range of values, the upper limit of Endosulfan, Lindane and DDT were much higher in our study. The results of Olutona and Aderemi [87] showed that Dieldrin, and Endosulfan, p,p’-DDT and p,p’-DDE concentrations were in the range of 0.99 – 11.01 mg/kg, 0.85 – 1.37 mg/kg, 1.05 – 1.82 mg/kg and 0.48 – 1.23 mg/kg, respectively in beans from markets in Ibadan metropolis. Meanwhile, in our study, Endosulfan was undetected, Dieldrin was within this range, p,p’-DDT and p,p’-DDE were much lower. Variations of this nature have been widely reported because the concentrations of pesticide residues in grains depend on several factors such merchant attitude and knowledge, type and quantity of pesticide used for preservation, duration of exposure of the grains etc.

## 5. Conclusion

One of the most crucial aspects of rural populations’ human capital in developing nations is health. A rising corpus of research demonstrates the benefits of good nutrition on productivity. However, the widespread usage of pesticides has resulted in numerous issues with human and environmental damage. Considering the numerous negative effects such substances have on society, it is vital to reevaluate their viability given the social and environmental issues associated with the usage of pesticides. To meet the need for food globally, alternative routes must be pursued to provide healthier food. Therefore, one of the primary findings of this study is that contamination is universal because no individual of any species throughout the entire basin appeared to be free of numerous pesticide residues in grains. The results frequently showed high inter- and intra-specific contamination, which makes sense given the target market and dietary diversity in the area. Therefore, efforts to locate and remove pollution sources will have an impact throughout Southwest Nigeria, not just on the markets that are previously recognized to be the most polluted. This study found that grains purchased from particular markets in southwest Nigeria contained numerous pesticide residues. The MRLs set by the EU or FAO/WHO or both were surpassed by 17 out of the total 27 pesticides reported in this work in at least one grain, despite the fact that there were no published codex MRLs for some pesticide residues in some grains. These residues were dispersed among the four classes of pesticides: carbamates, organochlorines, organophosphates, and pyrethroids. In actuality, 90% of the mainly banned organochlorine pesticides exceeded MRLs. The merchants who typically admit to the usage of pesticides were connected to their misuse; almost half of these merchants don’t even read the instructions on pesticide containers before using them. There is no doubt that the populous is exposed to the dangers brought on by eating wheat polluted with pesticides. While, cancer and physiological abnormalities are among the hazards. According to this research, the sale of beans, maize, millet, and rice grains in some markets in Southwest Nigeria may pose a serious health risk to the general public. In conclusion, routine assessments will give information on the contamination level in real time as well as the potential effects of unforeseen occurrences or subsequent actions controlling the flow of pollution. The sale of pesticides must be controlled in order to ensure that agricultural workers are trained in the safe use of these chemicals, and studies into the long-term effects of pesticide use on agricultural workers and the children who live in their homes are crucial as the volume of pesticide imports keeps rising. The Ministry of Health and Ministry of Agriculture employees can better target their educational efforts with the support of the information given above.

## Data Availability

All data produced in the present work are contained in the manuscript

## Ethics approval

This study entitled: “Evaluation of Pesticide Residues in Grain Sold at Selected Markets of Southwest Nigeria”, which was submitted for ethical approval on August 15, 2020 to the Research Ethics Committee of Kwara State University. For the development of this study, the fundamental ethical principles for research involving human beings, described and established by the Resolution 466/2012 and its complementary ones, was considered and approval was granted by the Kwara State University research ethics committee.

## Consent to participate

During the course of this research work, the participants were accorded the due respect so as to ensure co-operation and information collected were treated with utmost confidentiality. The cultures of the community were also respected during the course of the research work. Informed consent was obtained from all of the participants.

## Acknowledgements

We note with sadness the passing of our friend, colleague, and mentor **Prof. Mynepalli K. C. Sridhar**; working with him was wonderful and we strongly believe that he would have loved to coauthor this research paper – we miss him. We would like to say special thanks to all anonymous reviewer for providing very helpful comments for their valuable suggestions for improving this manuscript.

## Authorship contribution statement

**Modupe Abeke Oshatunberu:** Conceptualization, Investigation, Writing - original draft, Writing - review & editing. **Modupe Abeke Oshatunberu and Adebayo Oladimeji:** Investigation, Writing - original draft, Writing - review & editing. **Adebayo Oladimeji and Sawyerr Olawale Henry:** Resources, supervising, review & editing. **Modupe Abeke Oshatunberu, Morufu Olalekan Raimi:** Methodology, Project administration, Writing - original draft, Writing - review & editing. **Morufu Olalekan Raimi:** Data curation, Formal analysis, Writing - original draft, Writing - review & editing. **Modupe Abeke Oshatunberu, Adebayo Oladimeji, Sawyerr Olawale Henry:** Conceptualization, Data curation, Funding acquisition, Project administration, Writing - original draft, Writing - review & editing.

